# Non-CGG trinucleotide repeat expansions as pathogenic genetic mutations in Fragile X Syndrome

**DOI:** 10.1101/2025.10.22.25338597

**Authors:** Ana Carolina P. Cruz, Adriano Ferrasa, Isabeli do N. Masson, Janaina S. de Souza, Kelren S. Rodrigues, Roberto H. Herai

**Affiliations:** Laboratory of Bioinformatics and Neurogenetics (LaBiN), Experimental Multiuser Laboratory (LEM), Graduate Program in Health Sciences (PPGCS), School of Medicine and Life Science, Pontifícia Universidade Católica do Paraná (PUCPR), Curitiba, Paraná 80215-901, Brazil; Deparment of Informatics (DEINFO), Universidade Estadual de Ponta Grossa (UEPG), Ponta Grossa, Paraná 84030-900, Brazil; Departamento de Medicina, Disciplina de Endocrinologia Clínica, Universidade Federal de São Paulo, UNIFESP/EPM, São Paulo, São Paulo 04039-032, Brazil; Buko Kaesemodel Institute (ILK), Curitiba, Paraná 80240-000, Brazil

**Keywords:** *FMR1* gene, Fragile X syndrome, genetic variants, non-CGG repeats

## Abstract

**Purpose:** Fragile X syndrome (FXS) is a hereditary genetic condition, caused by the expansion of the trinucleotide CGG repeated over 200 times (full mutation) in the 5’UTR (untranslated region) regulatory region of the *FMR1* gene, which leads to the absence of FMRP protein. Although the clinical standard genetic confirmation for FXS diagnosis is limited to the repeats, the use of gene sequencing techniques allowed the identification of genetic variants that occur throughout the entire *FMR1* gene, including protein-coding and 3’UTR gene regions. These mutations may also cause the inactivation of *FMR1* gene, leading to the FXS phenotypes in individuals with CGG repeat expansions at a normal level (5-44 repeats) or at the premutation level (between 55 and 200 repeats), and not necessarily diagnosed with FXS.

**Methods:** To investigate how widespread the genetic mutations occurring throughout the *FMR1* gene locus are, we performed a Systematic Literature Review (SLR) to identify and synthesize a catalog of disease-causing mutations in the gene that are related to cause FXS or correlated conditions.

**Results:** After a detailed literature analysis, we found 44 single nucleotide variants (SNV) at the *locus* of the *FMR1* gene associated with developmental delay and/or intellectual disability, also including characteristics of FXS. Deletions involving the *FMR1* gene that remove several other genes were also found to be associated with FXS phenotype and ovarian problems, besides cases of mosaicisms with deletions and a case of germline mosaicism. Moreover, several of the mutations found, although occurring in distinct parts of FMR1 gene, can alter the aminoacid sequence of FMRP protein.

**Conclusion:** Our critical review presents several non-CGG repeat mutations in *FMR1* gene that are directly involved in the phenotypes found in Fragile X Syndrome, indicating that genetic screening for this neurodevelopmental condition should not be restricted to the CGG repeats.

## 1. Introduction

Fragile X syndrome (FXS) is a genetic condition that causes the most common form of inherited intellectual disabilities, with an estimated incidence of 1 in 7,000 men and 1 in 11,000 women (Crawford et al., 2001; Hunter et al., 2014). The symptoms presented by individuals with FXS are diverse, covering different degrees of cognitive impairment, in which are included intellectual defficiency, attention deficit hyperactivity, emotional disorders and autistic-like behavior (Bagni & Greenough, 2005). The physical characteristics such as narrow face, large ears, proeminent chin and large hands, strabismus, habit in shake arms and bite hands, are also observed symptoms in individuals with FXS (Carvalho, 2003; Hagerman et al., 1991; Quartier et al., 2017). Although there is no cure for FXS, it is known that its main cause is a mutation in the *FMR1* gene (Fragile X messenger ribonucleoprotein 1) (Bagni & Greenough, 2005).

*FMR1* is a protein coding gene, divided into 17 exons that covers 38 kb in the Xq27.3 *locus* (Eichler et al., 1993). This gene is transcribed in a 4,8 kb mRNA, which is used as a template to the synthesis of the FMRP (Fragile X messenger ribonucleoprotein 1 protein) (Bagni & Greenough, 2005).

Although, all the FMRP protein functions are still unclear, studies have shown that this protein is linked to RNA transport from nucleous to cytoplasm within a cell, and also in its own transportation to the ribosomes (Kim et al., 2009; Tabet et al., 2016). Thus, there are evidences that suggest that FMRP has an important role in regulating the translation of some mRNA, mainly in neurons dendrites and synapses (Serpa, 2008), in which FMRP is largely expressed (Bhakar et al., 2012; Darnell et al., 2011). Indeed, alterations in the metabolismo of mRNAs, important to the synapses structure and function, seems to be related to the learning and memory difficulties observed in subjects with FXS (Bagni & Greenough, 2005). In the central nervous system (CNS), it is estimated that aproximately 4% (Bhakar et al., 2012; Sethna et al., 2014) of total brain mRNA links to FMRP, which, in turn regulates its own translation into protein (Arsenault et al., 2016).

The main molecular basis of this syndrome is the expansion of the number of copies of CGG basis sequence (citosin-guanin-guanin) at 5’ end (5’UTR) to >200 repetitions (Mila et al., 2018; Roy Chowdhury et al., 2017; Zhou et al., 2016) (Fig. 1).

**Fig. 1.**
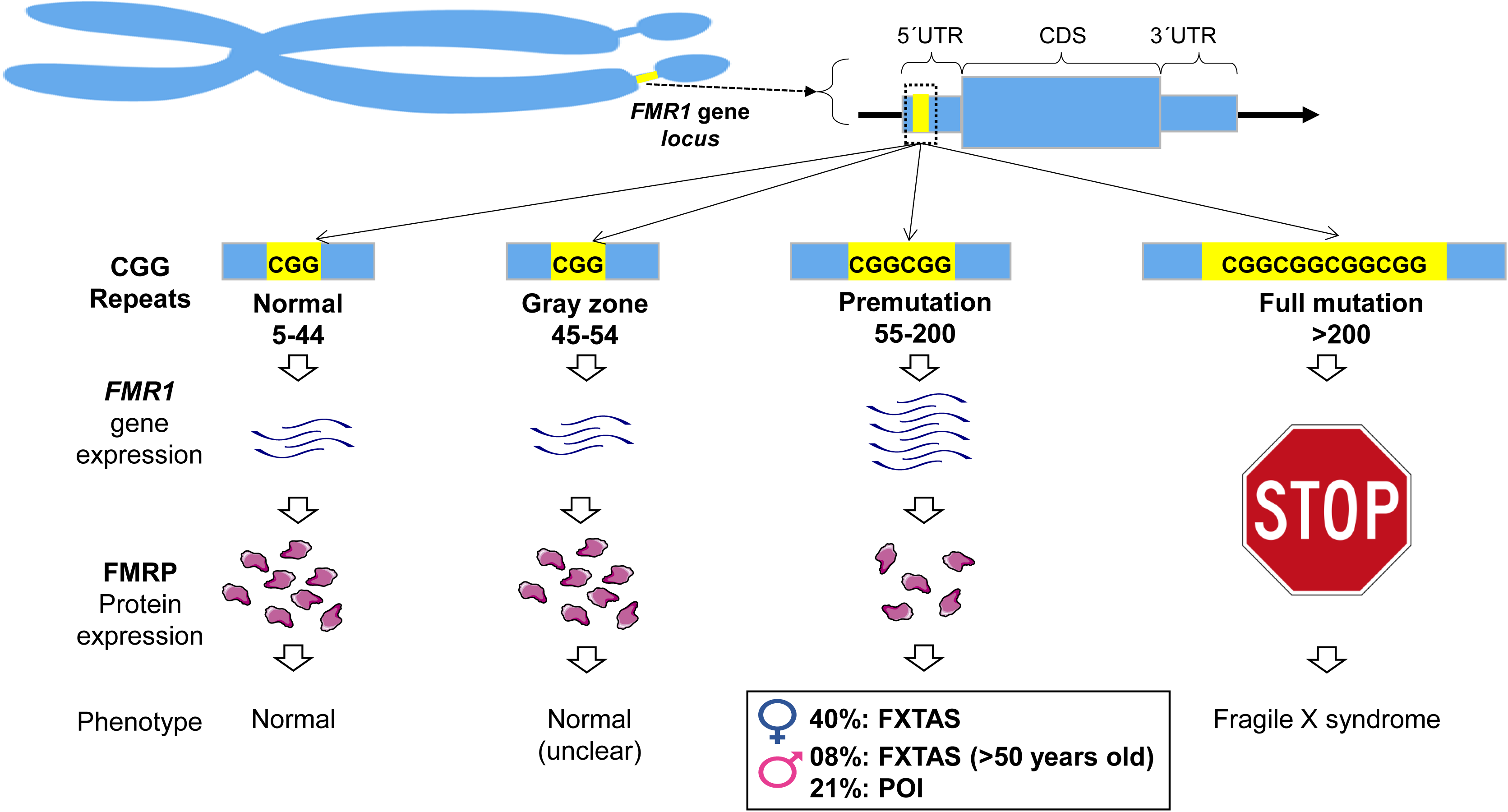
CGG trinucleotide repeat expansion that characterize Fragile X syndrome.

Since it is an unstable genetic mutation, the accumulation of mutations in the *FMR1* gene occurs in stages over the generations, in which the number of CGG repeats tend to expand (Fu et al., 1991; Macpherson & Murray, 2016). Depending on the number of repetitions and transmission instability, such sequence of trinucleotides may be classified in 4 allelic forms: normal (between 5 and 44 repetitions), intermediate or gray zone (GZ, between 45 and 54 repetitions), premutated (PM, 55 and 200 repetititions), and full mutation (FM, higher than 200 repetitions) (Chaudhary et al., 2014; McLennan et al., 2011).

In the full mutation, the CGG repetitions cause the hypermethylation of the *FMR1* gene and leads to transcriptional silencing of FMRP (Dean et al., 2016; Penagarikano et al., 2007). The presence of one allele with FM is sufficient to cause FXS, however the subject carriers of the PM allele do not present any phenotipc characteristic associated to FXS (Dean et al., 2016).

In this context, the rapid technological advances in the field of molecular genetics has contributed to the knowledge of several new genes and genetic factors involved in human diseases, including neurological conditions such as FXS (Durmaz et al., 2015). The use of techniques, in particular, genetic sequencing, has been increasinly helping the understanding of FXS. These techniques allow the identification of non-CGG related genetic variants that occur throught the *FMR1* gene *locus* and that may also cause its inactivation or alteration in the protein level, generating the FXS phenotype.

## 2. Material and Methods

The method used to perform this Systematic Literature Review (SLR) was based on the protocol developed by Kitcheham and Charters (2007). This review aimed to identify and synthesize the current knowledge about the variants that were reported in the *FMR1* gene and its association with the FXS phenotype.

To distinguish the articles that identified and/or studied the genetic variants of the *FMR1* gene, the SLR was done on PubMed, Scopus, and Bibliotec Virtual em Saúde (BVS) databases untilOctober XX, 2025.

To include all the existing studies regarding the subject considered, the following terms were related: gene/protein, genetic variants, and molecule, resulting in a combination of 51 keywords for the construction of the search string (Table S1).

The results obtained from the electronic search were extracted directly to a spreadsheet and tabulated with the following information: Author, title, year, journal, volume, edition, DOI, link, and database. Next, a triage was carried out to remove duplicate articles and then to select relevant articles for this SLR.

After the duplicate papers were removed, two authors reviewed each title and abstract. These preliminary lists of selected papers were then analyzed for their full content to identify only the studies that met the inclusion criteria.

Studies that were included in the revision were the ones that showed different genetic variants of CGG repeats, epigenetic data at the locus of the human *FMR1* gene, or any organism that presents an *FMR1* homologue.

We excluded studies that involved variants in other genes, functional aspects of the FMRP protein, therapeutic targets, and diagnostic techniques.

At the end of this stage, a complete list presenting the rejected studies and the reason for the rejection was kept. The selected articles were analyzed to collect the following information: variant, variant type, altered allele, reference allele, substituted amino acid position, inheritance standard, position in the genome (standardized to the genome assembly GRCh38/hg38), isoform, gene region, variant effect, individual phenotype/condition.

## 3. Results and Discussion

Based on the defined criteria for this SLR, which followed a protocol defined by Kitchenham and Charters (2007), a total of 6,121 articles were extracted from the PubMed, Scopus, and HVL search databases. In this review, we considered studies published untilOctober XX, 2025. After the removal of duplicate papers and analysis of their full-text content, and applying the defined inclusion/exclusion criteria, there were 56 papers left that were used to collect the information for this SLR (Fig. 2).

**Fig. 2.**
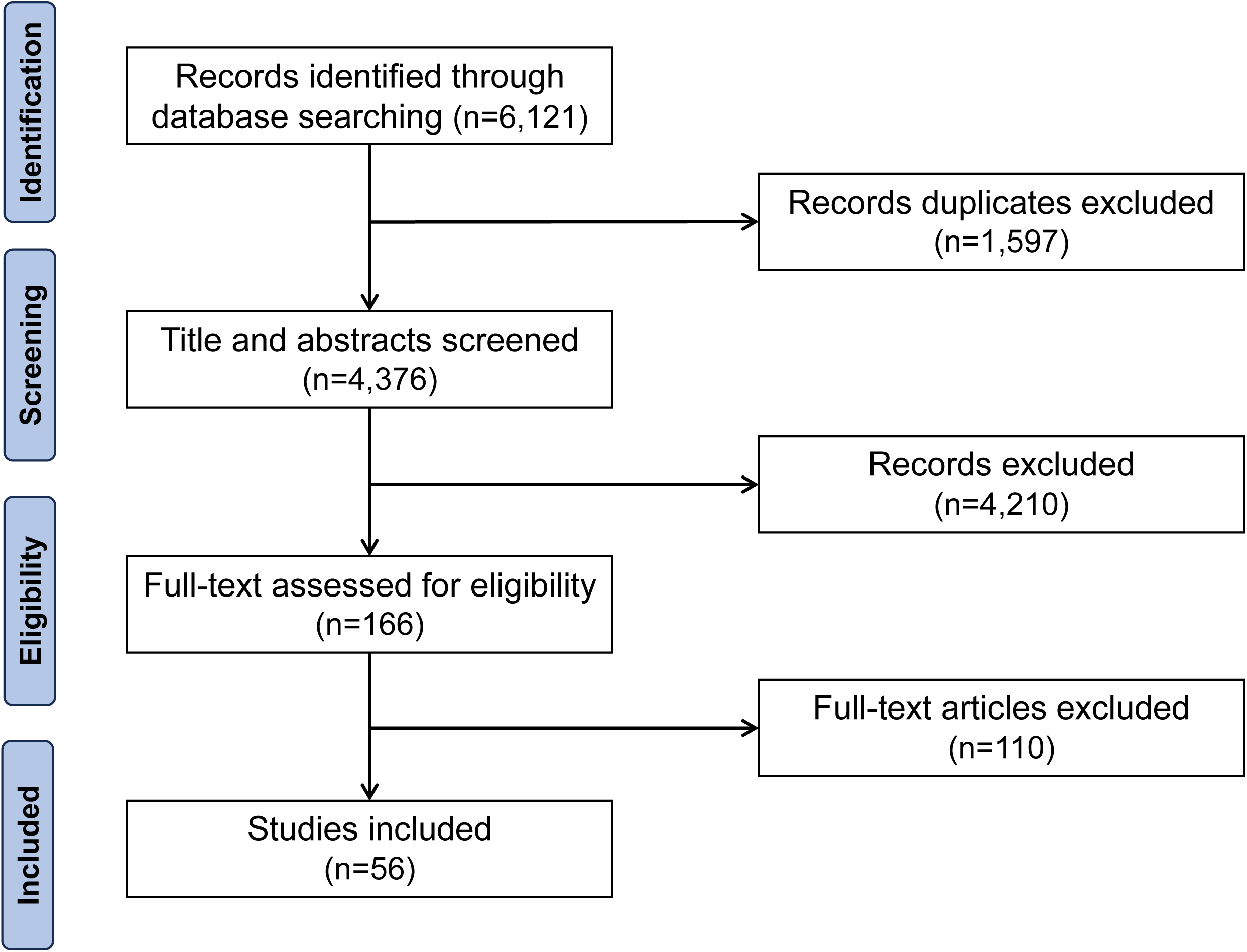
Flowchart for identification and selection of articles from the studies included in the Systematic Review of Literature. The diagram details the number of studies found and selected after applying the inclusion and exclusion criteria at each step of the literature search

### 3.1. Single nucleotide variants (SNV)

From several studies based on the use of sequencing techniques, it was possible to detect 44 SNV (Fig. 3) at the *locus* of the *FMR1* gene. Among these variants reported, carrier individuals presented a broad spectrum of clinical phenotypes: 34 of these variants (Table S2) were associated with developmental delay and/or intellectual disability, although general symptoms are associated with FXS, and 10 variants were associated with clinical characteristics found in the syndrome (Table S3).

**Fig. 3.**
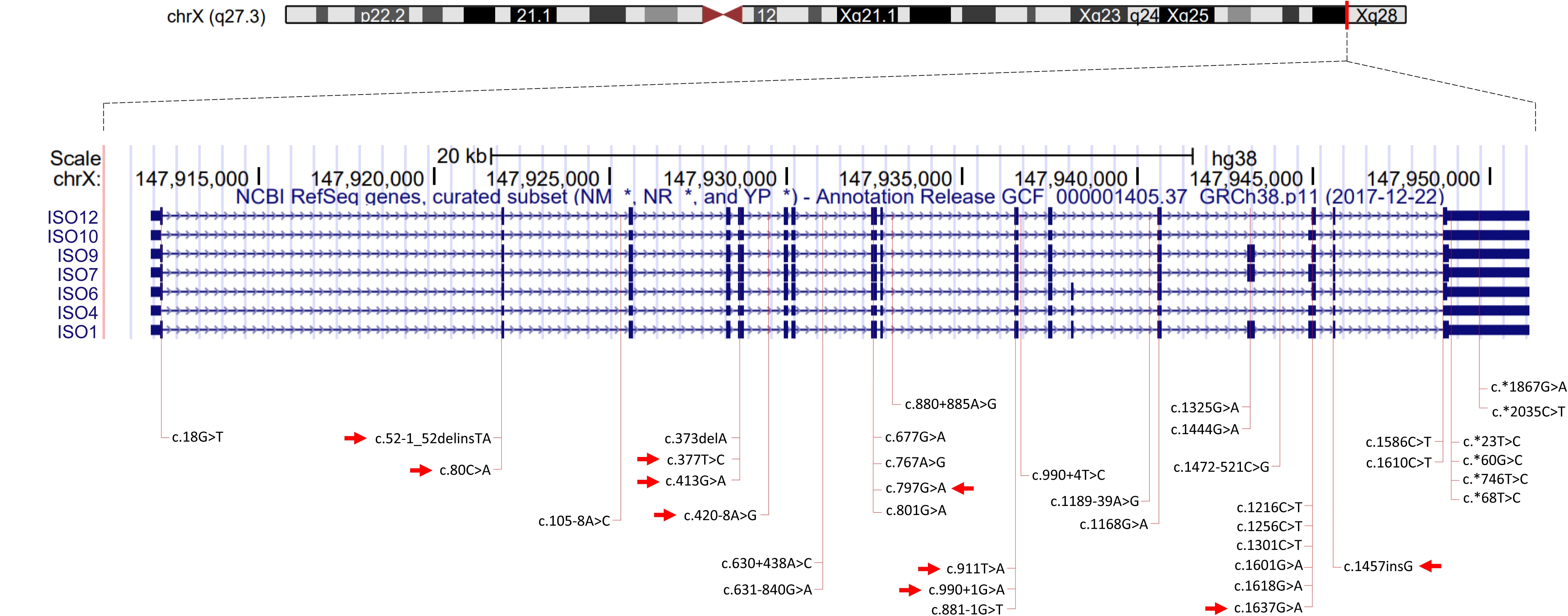
*Locus* of the *FMR1* gene indicating the point genetic variants identified by the RSL. All variants are listed in the exons and intons of the *FMR1* gene (chrX:147,911,951-147,951,127) in the human reference genome GRCh38, with 39,177 bases of length. Variants occurring in the promoter region of the gene are not shown in the figure (c.−413C>G, c.−332G>C, c.−293T>C e c.−254A>G). The highlighted variants with red arrows represent those found in individuals with phenotype associated with FXS.

Five variants were reported in patients who presented clinical signs of intellectual disability and developmental delay, but with normal CGG repeat expansions in *FMR1* (Handt et al., 2014). These mutations include two in the coding region, two in the 3’UTR region (c.*60G>C; c.*68T>C), and one in an intronic region (c.1189-39A>G). The variants found in the non-coding regions do not have a predicted pathogenic effect. On the other hand, both missense mutations, c.1444G>A e c.1601G>A, were predicted as pathogenic. In the variant c.1444G>A, the substitution of the nucleotide G by A, altered the aminoacid glycine by serine in the position 482 of protein (p.Gli482Ser), compromising the interaction of FMRP with other proteins (such as 58 KDa Microspherule Protein - MSP-58, Survival Motor Neuron Protein - SMN and RAN Binding Protein 9 - RANBP9). The substitution of c.1601G>A, in turn, influences the FMRP binding with RNA and polyribosomes. This occurs since the change of arginine by a histidine in the 534 position (p.Arg534His) occurs in the RGG box protein domais, a region rich in arginin and glicin involved in binding RNA.

In another large investigation, it was analyzed a cohort of 963 male patients with a general diagnosis of developmental delay and normal CGG repeat expansion (Collins et al., 2010), found the variants c.18G>T (p.Val=) and c.413G>A in the *FMR1* gene *locus*. In the second SNV mutation, the exchange of guanine by an adenine caused an arginine substitution for glutamine at position 138 (p.Arg138Gln) in the nuclear localization signal (NLS) of the FMRP protein. In the same study, another 10 variants in non-coding regions but having a significance impact in the protein function were detected as follows: c.105-8A>C, c.630+438A>C, c.631-840G>A, c.880+885A>G, c.990+4T>C, c.1472-521C>G, c.*23T>C, c.*2035C>T, c.*1867G>A and c.*746T>C. One of these reported variants, the c.*746T>C, was also reported by an independent study (Suhl et al., 2015), in a patient diagnosed with autism spectrum disorder, attention deficit hyperactivity disorder (ADHD), and moderate intellectual disability (intelligence quotient, IQ= 47). This variant found in the 3’UTR region of the gene *FMR1*, is responsible for the interruption of RNA-protein interaction, eliminating the normal response to glutamate signaling in cultured primary neurons and reducing FMRP levels. The half-brother of the patient, with ADHD and IQ = 67, has the same genetic variant, suggesting this variant has low penetrance.

Another study used microarray and exome sequencing of samples from children with undiagnosed developmental disorders and their corresponding parents. After genetic data analysis, it was found several genetic variants, and two were present in the *FMR1* gene: c.377T>C and c.1325 G>A (Wright et al., 2015). Both missense variants occur in the coding region of the gene. The first is responsible by the change of fenilanine by a serine at position 126 (p.Phe126Ser), and the second variant, an arginine is replaced by glutamine at position 442 (p.Arg442Gln).

Similarly, almost one thousand individuals with moderate to severe intellectual disabilities were analyzed by next-generation sequencing (NGS) methodology (Grozeva et al., 2015). In *FMR1*, 6 missense variants were found in ten different individuals: c.1301C>T (p.Ala434Val), c.767A>G (p.Asp256Gli), c.677G>A (p.Arg266Lis), c.1586C>T (p.Tre529Ile), c.1168G>A (p.Ala390Tre) and c.1610C>T (p.Ser537Leu). In another case of intellectual disability, in exon 15 of *FMR1*, the SNV variant c.1618G>A (p.Gly540Glu) was found (Hu et al., 2016).

SNVs in the promoter region of the *FMR1* gene were also identified. Collins and collaborators (2010) found three cases of this variant: c.−332G>C, c.−293T>C, and c.−254A>G (Collins et al., 2010). These genetic variants decrease the gene expression and may hinder the onset of *FMR1* transcription, also resulting in reduced levels of functional FMRP (Suhl & Warren, 2015). Another study showed the segregation of the variant c.−413C>G (Grasso et al., 2010) in three generations of the same family (segregated in healthy maternal grandfather, healthy mother and son with mild intellectual disability). This same variant has previously been reported in another patient with an unknown origin of the presented intellectual disability (Milà et al., 2000).

The above-cited cases encompass symptoms that, although of a broad spectrum, they are associated with FXS. Other cases demonstrate an association between variants in the *FMR1* gene with typical characteristics of FXS. In a male patient with intellectual disability, developmental delay, and convulsions (characteristics present in 10-20% of patients with FXS) and with CGG repetitions at the intermediate level (45 repetitions), it was found the missense variant c.413G>A (p.Arg138Gln) with a maternal heritage (Collins et al., 2010; Myrick et al., 2015). Another male subject carrying this same SNV mutation had developmental delays, autistic behavior, attention deficit hyperactivity disorder, seizures, handshake, elongated face, large ears, soft hands, hyperextensible fingers and flat feet (Sitzmann et al., 2018). It was reported that this variant lead to a partial loss of the FMRP protein function (Myrick et al., 2015; Suhl & Warren, 2015). The study also showed that the abilities of FMRP to bind to mRNAs in the post-synaptic translation are preserved, however, the protein becomes unable to maintain its presynaptic structural effects. This occurs because the variant severely impairs the interaction of FMRP with the BK channels (K^+^ channel of high conductance activated by Ca^2+^), an important structure in the regulation of neurotransmitter release and neuronal excitability.

Similarly, another work (Okray et al., 2015) found in a male subject with FXS typical clinical phenotype (intellectual disability, autistic behavior, impaired social interaction) and a normal expansion of 41 CGG repeats, the variant c.1457insG. The insertion of a single guanine caused a frameshift that adds a new peptide sequence of 22 amino acids followed by a premature stop codon, resulting in a truncated protein (p.Gly538fs*23).

Another typical individual with FXS phenotype (developmental delay, elongated face, large ears, prominent mandible, and macroorchidism) was associated with the *de novo* SNV variant c.911T>A (De Boulle et al., 1993). The variant is responsible for the exchange of the amino acid isoleucine by asparagine (p.Ile304Asn) located in the KH2 domain of the FMRP protein. Using Molecular Dynamics Simulation Techniques (Di Marino et al., 2014), it was shown that this missense variant destabilizes the hydrophobic nucleus of the protein, causing malfunctions and potentially leading to the observed phenotypes.

Two other cases of male patients with classic FXS characteristics (Lugenbeel et al., 1995) presented two different variants. These mutations are a *de novo* deletion (c.373delA; p.Thr125Leufs*35), found in a young patient, and the second mutation is a hereditary alteration of two base pairs in an adult (c.52-1_52delinsTA), both responsible by one truncated change in the protein aminoacid sequences.

A missense variant, of maternal inheritance, was found in an individual with typical FXS physical behaviors and characteristics. The patient had 23 CGG repetitions (normal), but on exon 8 it was found the SNV variant c.797G>A (Myrick et al., 2014). This amino acid exchange (p.Gly266Glu) interrupts the RNA-binding protein function, no longer allowing the regulation of translation by FMRP. This occurs because structurally, at position 266, the protein probably requires a small, non-polar, flexible amino acid, characteristic of glycine rather than glutamate.

Another study found two mutations in two distinct individuals: c.990+1G>A and c.420-8A>G (Quartier et al., 2017). Despite presenting CGG repetitions within the normal level, the patients presented characteristics consistent with FXS. The variant c.990+1G>A, causes aberrant splicing in the transcript, leading to the loss of a 110 pb in the exon 10, creating a frameshift after amino acid 294, as a consequence, the protein has a premature stop codon (p.Lys295Asnfs*11). The resulting truncated FMRP does not have essential domains to the normal function, such as nuclear export signaling and RGG box domains. However, c.420-A8>G is an intrinsic variant that leads to the insertion of 5 nucleotides in the intron 5. The result is similar to the previous mutation: a truncated protein lacking essential domains caused by a frameshift and by the emerge of a premature stop codon (p.Met140Ilefs*3).

A nonsense variant c.80C>A (Gronskov et al., 2011), that predicts the change of serine by a premature stop codon (p.Ser27*), was found in a 35 year old man and in his heterozygous mother to the variant. The patient’s symptoms included: intellectual dysfunction, facial dysmorphism, macroorchidism, epilepsy, autistic characteristics and little use of language, and classic clinical FXS symptoms.

Wang and collaborators (1997) found a substitution of the nucleotide C by T in intron 10 causing an alternative splicing that excludes exon 10. Deletion of exon 10 results in a frameshift and a premature termination of the translation, which removes the highly conserved region encoding the KH2 and RGG box protein domains. In the same individual, another mutation was found on exon 15 c.1637G>A (Quartier et al., 2017), which causes the substitution of arginine by histidine at amino acid position 546 (Arg546His) of FMRP. However, as the mutation in the intron produced a truncated FMRP, the mutation c.1637G>A would not have meaning in the formation of the disease. Thus, the mutation in the intro 10 would be sufficient for the FXS development (Wang et al., 1997).

After combining and comparing all the SNV mutations found in this review, were reported three stopgain variants in the *FMR1* gene (c.52-1_52delinsTA, c.80C>A, c.1216C>T), 4 frameshift variants (c.373delA, c.420-8A>G, c.990+1G>A e c.1457insG), 18 nonsynonymous variants (c.377T>C, c.413G>A, c.677G>A, c.767A>G, c.1168G>A, c.1325G>A, c.1444G>A, c.1601G>A, c.1618G>A, c.1301C>T, c.1586C>T, c.1610C>T, c.797G>A, c.911T>A, c.1637G>A, c.1256C>T, c.881-1G > T, c.801G > A) (Fig. 4), one synonymous variant (c.18G>T) and 17 in non-coding regions of the *FMR1* gene (c.−413C>G, c.−332G>C, c.−293T>C, c.−254A>G, c.105-8A>C, c.630+438A>C, c.631-840G>A, c.880+885A>G, c.990+4T>C, c.1189-39A>G, c.1472-521C>G, c.*23T>C, c.*60G > C, c.*746T>C, c.*68T > C, c.*1867G>A, c.*2035C>T) (Table 1).

**Fig. 4.**
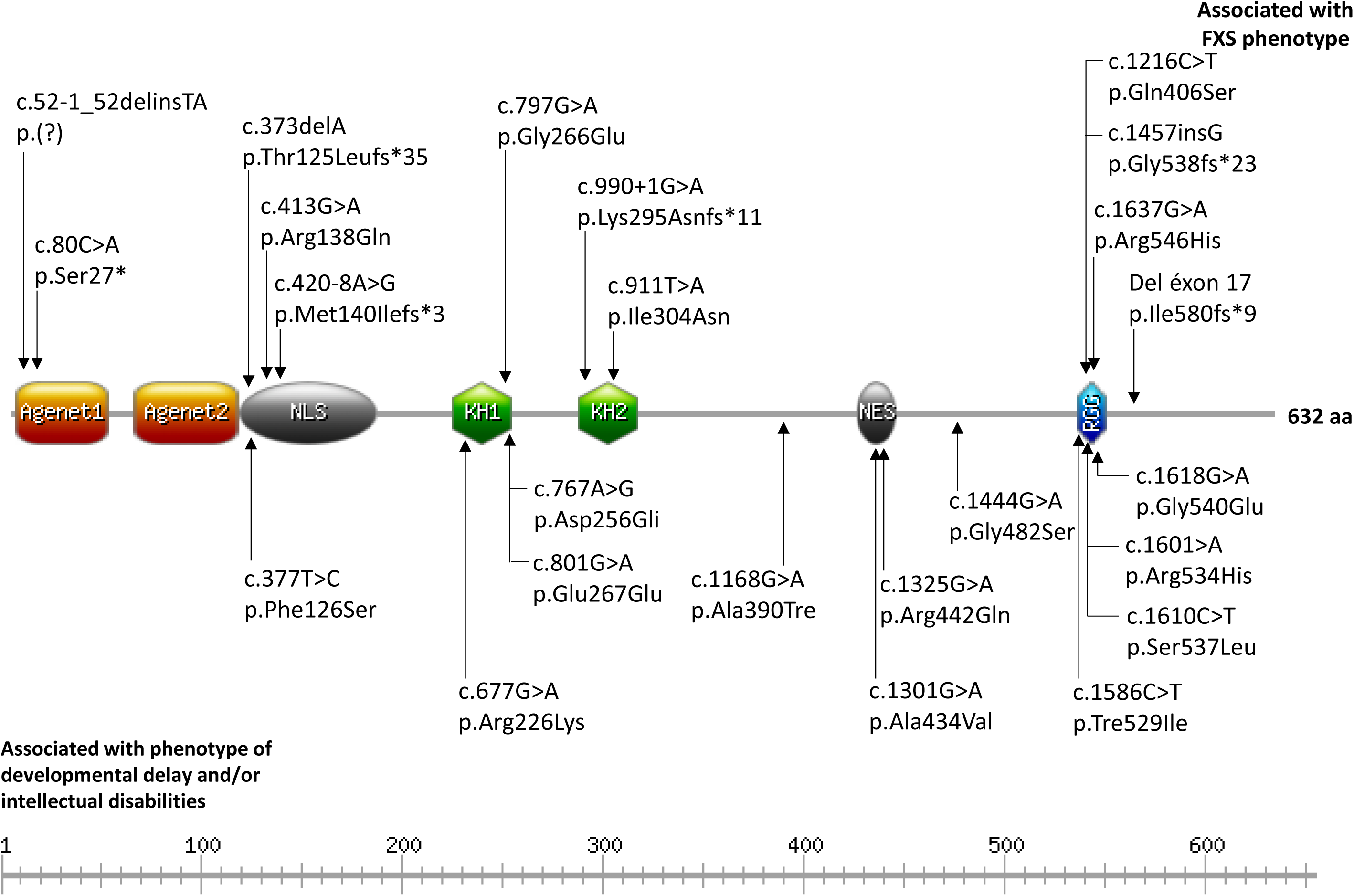
Location of genetic variants found along FMRP protein in individuals with FXS or with related. The variants indicated at the top of the FMRP are those found in individuals with typical FXS phenotype. The variants listed below the FMRP are those found in individuals who had intellectual disability and/or developmental delay. Abreviations: Agenet, Agenet-like domains; KH, K homology domain; NES, nuclear export signal; NLS, nuclear localization signal; RGG, glycine–arginine rich domain (RGG-box).

Finally, we also found a case of a 17-year-old boy with a milder phenotype, including the lack of typical facial dysmorphism. Interestingly, this patient had FM of approximately 250 CGG repeats, but the gene was partially methylated (Haberlandt et al., 2014). This epigenetic pattern with a partially methylated FM showed lesser intellectual impairment and milder facial dysmorphism when compared to individuals with FM and complete gene methylation.

### 3.2. Deletions

The deletions found involving the *FMR1* gene (Fig. 5) can be divided into two classes: the first those that remove several other genes including the *FMR1* (Fig. 5B) and the second those that occurred only at the *locus* of the *FMR1* gene (Fig. 5C).

**Fig. 5.**
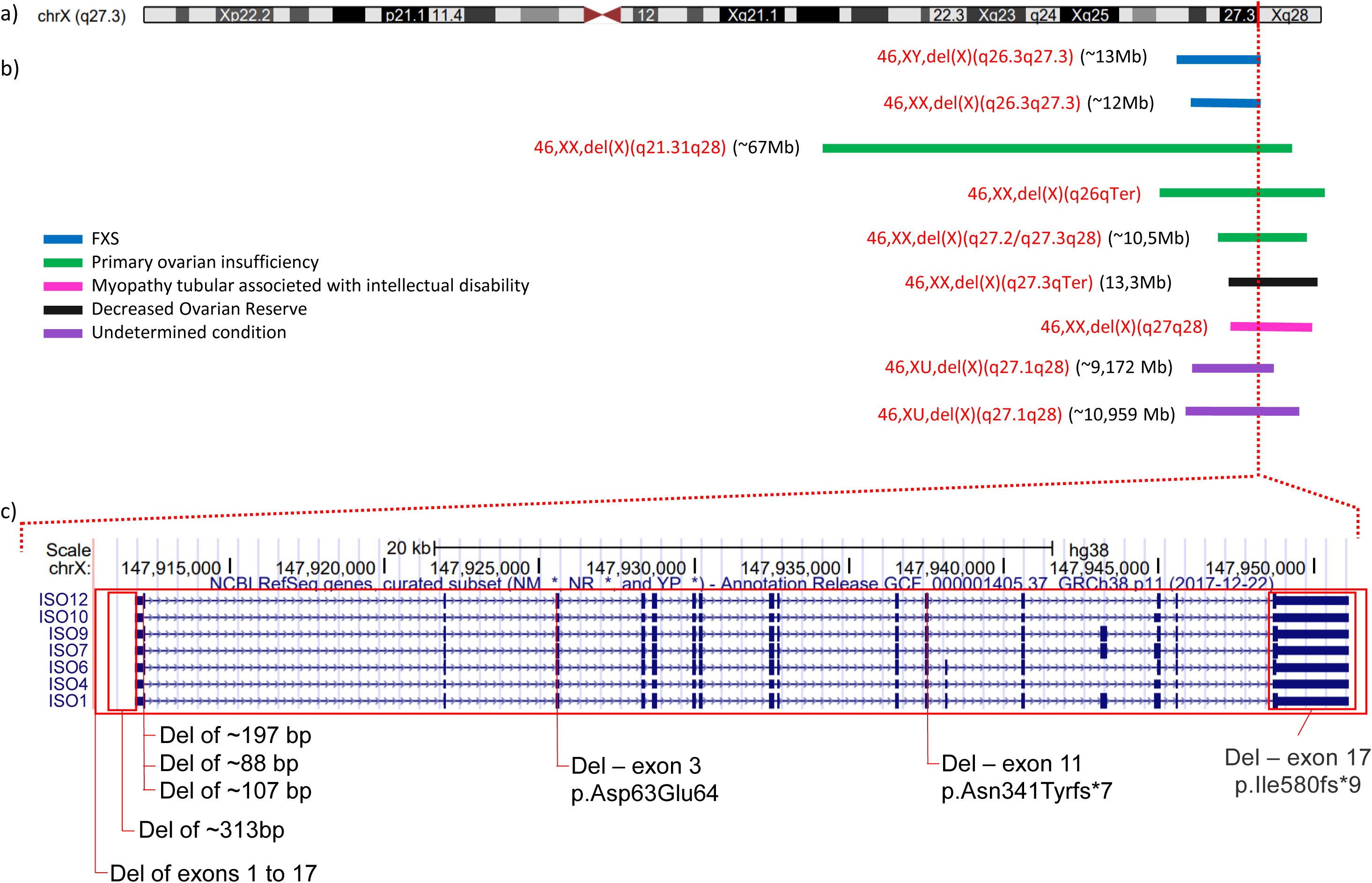
*Locus* of the *FMR1* gene indicating the deletions identified by the RSL. **A:** Representation of the X chromosome. The dotted red line represents the position of the *FMR1* gene (chrX: q27.3). **B:** localization of 9 deletions responsible for the elimination of several genes including *FMR1*. The dotted red line represents the position of the *FMR1* gene; **C:** localization of8 deletions that occurred in the *FMR1* gene.

In the second class (Fig. 5C), there is a case of a small deletion, with approximately 197 bp, that was found in a patient presenting a typical FXS phenotype (intellectual disability, repetitive speech, macroorchidism, prominent jaw, large ears, flat feet, ADHD, poor eye contact, convulsions, and autistic behavior) (Viveiros, 2013). The deletion in exon 1 involves part of the CGG repeat region at 5’UTR and its proximal region.

Another deletion was found in three siblings with fragile X features (Quartier et al., 2017) and, in a heterozygous state, in an asymptomatic mother. This deletion removes exon 17 from the *FMR1* gene and its acceptor splice site on the *FMR1* gene (hg19 - NM_002024: g.(?_ 147030202)_(147046357 _?)del) and leads to a truncated protein (p.Ile580fs*9).

In one of the cases analyzed, the subject of study presented intellectual disabilities associated with a “particular phenotype”, but not exactly the ones commonly seen in FXS individuals. He has a 60 kb hemizygous deletion on Xq27.3, which encompasses and deleted the entire *FMR1* gene within the boundaries at 146,990,647 and 147,058,715 bp. Curiously, his mother had the same deletion in heterozygous state, but was a phenotypically and cognitively normal woman (Gómez-Rodríguez et al., 2022).

There was found a microdeletion paired with mosaic premutation in an individual with a mother with large CGG repeats and a full-mutation sibling. The subject had a 313 bp deletion in the promoter region of *FMR1* upstream of the CGG repeats and presented with mild FXS features (Hnoonual et al., 2024)

Among the deletions that remove several genes (Fig. 5B), we found the deletion including Xq26.3-Xq27.3 (13 Mb) in a male patient with intellectual disability and facial features consistent with FXS (Wolff et al., 1997). In the same region (Xq26.3-Xq27.3) a *de novo* 12 Mb deletion was reported in a woman with moderate to severe intellectual disability, long and narrow face, prominent chin, and large ears (Wolff et al., 1997).

Another associated disease with large deletions found in this SLR is premature ovarian failure (POF). This disease is also associated with PM, occurring in approximately 21% of premutated women (Biancalana et al., 2015; Dean et al., 2016). But in the cases discussed below, the causes of the disease are large deletions involving several genes, among them *FMR1*.

A 67.355 Mb (Xq21.31-Xq28) *de novo* deletion occurred only in one of the X chromosome in a POF female (Beke et al., 2013). The other X chromosome, normal and activated, did not present the *FMR1* methylated allele. This region is composed of 795 genes, of which, 10 were already associated with POF, including *FMR1.* Another *de novo* deletion that has also included *FMR1*, occurred in the cytoband Xq26-Xqter (Ferreira et al., 2010). Another deletion of maternal inheritance, with an average length of 10.5 Mb (Xq27.2/Xq27.3-Xq28) (Eggermann et al., 2005), was found in a woman with POF who presented mild stigmata of Turner Syndrome. This woman inherited from her father the normal *FMR1* allele (31 CGG repetitions) and the deleted allele came from her mother.

In another interesting case, it was found the condition Decreased ovarian reserve (DOR), a primary infertility disorder characterized by a reduction in the number or quality of oocytes. In this case, it was found a genomic translocation that involved the chromosomes X and 18 (der(X)t(X;18)(q27;q22)). The genetic analysis defined a subtelomeric deletion spanning 13.3 Mb of Xq27.3-Xqter, and a doubling spanning 13.4 Mb, from 18q22.1-18qter (Fusco et al., 2011)(Fusco et al., 2011).

Finally, another condition known as myotubular myopathy and associated with intellectual disability was found in a woman with a *de novo* heterozygous deletion between the chromosomal bands Xq27-Xq28 (Dahl et al., 1995). The authors suggest that the developmental delay, particularly the greater delay in language, as observed in the patient, may be a manifestation of FXS since the *FMR1* gene is included in the deleted region. However, since there are other genes in the region of this deletion, these may also play a role in the phenotype.

### 3.3. Mosaicism

Another type of variant found by this review corresponds to genetic mosaicism. Genetic mosaicisms are characterized by the presence of different sizes of CGG repeats between different cells. The mosaicisms for FM and PM are relatively common (Biancalana et al., 2015)(Biancalana et al., 2015). Here we describe unusual cases of mosaicism, which in addition to alleles for FM and PM, present deletions, and a case of germline mosaicism (Table 2).

One of these cases involves germline mosaicism of a 300 kb deletion that spans the entire *FMR1* gene (NC_000006.12:g.146414070_146715778del) (Jiraanont et al., 2016)(Jiraanont et al., 2016). This deletion was found in a male patient and his neurotypical mother.

In another study, it was found an individual with genetic mosaicism, with part of his lymphocyte cells having a normal allele with 23 CGG repeats, and another part of cells with a ∼1Mb (1,013,395pb) deletion, corresponding to approximately 90% of cells. The mosaic deletion is responsible for the removal of the whole *FMR1* gene and its neighbor *FMR1NB* (*FMR1* Neighbor) gene (Coffee et al., 2008)(Coffee et al., 2008).

In another case, it was found mosaicism that included methylation of an expanded allele and a *de novo* microdeletion of ∼80bp in the *FMR1* promoter. This patient was diagnosed with FXS and had compatible clinical features with FXTAS that manifested around 30 years of age (Hwang et al., 2016)(Hwang et al., 2016).

In a classical FXS family patient, but without the characteristic FXS phenotype, including an unexpected IQ within the considered normal level, mosaicism was also found in two cell types (Govaerts et al., 2007)(Govaerts et al., 2007). The patient presented a FM mosaic pattern with alleles of 230 and 450 CGG repeats (60%), PM (170 repeats, 25%), and a deletion of 175bp (15%) in the *FMR1* gene. This deletion removed the CGG repeat along with 61 bp before the CGG repeat and 27 bp after.

In another FXS family, it was found a diverse mosaicism case, involving a methylated FM (10%), unmethylated FM/PM (37%), and a small deletion (53%) (Han et al., 2006)(Han et al., 2006). In this patient, FMRP was expressed in only 22% of his cells and the mRNA level was increased by 3.6 fold. The low level of FMRP expression is attributed to the deletion spanning the entire region of the CGG repeats plus 42bp upstream thereof. The high levels of mRNA, therefore, are associated with the PM that the patient carries. The patient still has a brother with classical methylated FM and a half-brother who also presents mosaicism but based on the methylation level of the FM regions, with total methylation (55%) and unmethylated repeats (45%).

In another case, it was found a mosaicism with a methylated FM (300 to 350 CGG replicates) and a 905 bp deletion (Arocena et al., 2000)(Arocena et al., 2000). This deletion encompasses the CGG repeats and excludes a start codon (ATG) of the *FMR1* gene, leading to the absence of FMRP in the individual, since neither the FM allele nor the deletion could be transcribed. Another case involving the methylation state and deletion was detected in a male with ADHD, anxiety, finger biting, poor eye contact, hand flapping, long face, and prominent ears (Jiraanont et al., 2017)(Jiraanont et al., 2017). He has the presence of expanded alleles (methylated and unmethylated full mutation alleles) with an allele encompassing a deletion across the CGG repeat that was present in 13% of the cells while cells carrying hypermethylated full mutation alleles were in approximately 60% of the cells.

In an elder individual, a 68 year-old man, it was found a mosaicism for FM and a 100 bp deletion from the locus of the *FMR1* CGG repeats (de Graaff et al., 1996)(de Graaff et al., 1996). This deletion did not impair the transcription and translation of FMRP, which was present in 28% of the lymphocytes of the patient who had a clinical phenotype of FXS.

Another peculiar case involves a 70-year-old man who had two PM daughters and three grandchildren with FXS. He had no ID and had a normal cognitive function. Only after the age of 64, he began to present late-onset of neurological symptoms of FXTAS (Santa María et al., 2014)(Santa María et al., 2014). It was observed that the *FMR1* mRNA level was 7 fold above the normal mean and that FMRP expression levels were reduced (38% of normal). This phenotype is the result of a mosaic of unmethylated FM size (with CGG repeat sizes ranging from 180 to 410).

An intriguing case was presented in the study of Winarni et al. (2025). They described a family with five adopted children, in which two of them had full-mutations of *FMR1*, the two identical-twin triplets had mosaic full-mutations paired with a 107 bp deletion and the other sibling was normal. Methylation was observed in 70% and 60% of the cells of the two identical-twin triplets (Winarni et al., 2025).

### 3.4. Epigenetic modificattions

Epigenetics refers to DNA changes that do not involve changes in the sequence itself (Francis, 2015)(Francis, 2015). Epigenetic silencing of genes is seen in several diseases caused by trinucleotide replicates (Colak et al., 2014; Nageshwaran & Festenstein, 2015)(Colak et al., 2014; Nageshwaran & Festenstein, 2015). Some epigenetic mechanisms regulate part of DNA transcription such as the formation of heterochromatin and associated histone modifications, DNA methylation, antisense transcription (3’→5’), and the formation of complex DNA structures (DNA-RNA hybrids, RNA and DNA tríplex, and RNA loops) (Nageshwaran & Festenstein, 2015)(Nageshwaran & Festenstein, 2015)(Nageshwaran & Festenstein, 2015)(Nageshwaran & Festenstein, 2015).

In this review, it was found distinct types of epigenetic modifications associated with FXS. They are briefly discussed in the following topics to highlight how they can be formed and causes the phenotypes found in FXS.

#### 3.4.1. Epigenetic effects mediated by CGG repeats

Errors in replication and DNA repair pathways appear to lead to the instability of CGG repeats (Usdin & Kumari, 2015)(Usdin & Kumari, 2015)(Usdin & Kumari, 2015)(Usdin & Kumari, 2015). Studies with human embryonic stem cells (hESCs) show that changes in the replication fork progression occur due to the absence of initiation sites at the *FMR1 locus*(Gerhardt, 2017)(Gerhardt, 2017)(Gerhardt, 2017)(Gerhardt, 2017).

In addition, CGG repeats that are present in the *FMR1* gene and the transcript may form secondary structures (Usdin & Kumari, 2015)(Usdin & Kumari, 2015)(Usdin & Kumari, 2015)(Usdin & Kumari, 2015), which appear to hinder the progression of DNA polymerase during replication (Gerhardt, 2017)(Gerhardt, 2017)(Gerhardt, 2017)(Gerhardt, 2017). Examples of secondary structures formed from the repeats are hairpin, R-loop and duplex RNA-DNA (Colak et al., 2014; Nageshwaran & Festenstein, 2015; Usdin & Kumari, 2015)(Colak et al., 2014; Nageshwaran & Festenstein, 2015; Usdin & Kumari, 2015)(Colak et al., 2014; Nageshwaran & Festenstein, 2015; Usdin & Kumari, 2015)(Colak et al., 2014; Nageshwaran & Festenstein, 2015; Usdin & Kumari, 2015).

In individuals with stable sequences, the presence of the AGG trinucleotide interrupting the CGG repeats decreases the stability of these secondary structures. Thus, the AGG interrupts prevent the expansion of CGG replicates (Jarem et al., 2010; Usdin et al., 2014)(Jarem et al., 2010; Usdin et al., 2014)(Jarem et al., 2010; Usdin et al., 2014)(Jarem et al., 2010; Usdin et al., 2014).

#### 3.4.2. Epigenetic effects on premutation (PM)

In PM, CGG repeats lead to increased initiation of *FMR1* transcription (Kraan et al., 2018)(Kraan et al., 2018)(Kraan et al., 2018)(Kraan et al., 2018). Thus, premutated individuals have 2 to 10 times more mRNA than individuals with CGG replicates at a normal level (Eslami et al., 2018; Sheridan et al., 2011; Usdin & Kumari, 2015)(Eslami et al., 2018; Sheridan et al., 2011; Usdin & Kumari, 2015)(Eslami et al., 2018; Sheridan et al., 2011; Usdin & Kumari, 2015)(Eslami et al., 2018; Sheridan et al., 2011; Usdin & Kumari, 2015), but without a subsequent increase of FMRP protein indicating a gain mechanism of toxic RNA function (Nageshwaran & Festenstein, 2015)(Nageshwaran & Festenstein, 2015)(Nageshwaran & Festenstein, 2015)(Nageshwaran & Festenstein, 2015). Active chromatin marks are found at the 5’ UTR end of *FMR1* in PM carriers (Usdin & Kumari, 2015)(Usdin & Kumari, 2015)(Usdin & Kumari, 2015)(Usdin & Kumari, 2015) and increased transcription is associated with an increased abundance of acetylated histones in the *FMR1* promoter (Usdin et al., 2014)(Usdin et al., 2014)(Usdin et al., 2014)(Usdin et al., 2014).

*In vitro,* long traces of CGG repeats exclude nucleosomes, which may increase the accessibility of transcription factors in the promoter (Usdin et al., 2014; Usdin & Kumari, 2015)(Usdin et al., 2014; Usdin & Kumari, 2015)(Usdin et al., 2014; Usdin & Kumari, 2015)(Usdin et al., 2014; Usdin & Kumari, 2015).

In addition to the higher density of CpG islands within CGG repeats, other factors may also contribute to the overexpression of PM alleles. For example, the ATRX protein (ATRX, chromatin remodeler - member of the SNF2 family of helicases/ATPases) facilitates the lengthening of transcription through G-rich regions (such as the *FMR1 locus*), reducing transcription block (Usdin & Kumari, 2015)(Usdin & Kumari, 2015)(Usdin & Kumari, 2015)(Usdin & Kumari, 2015).

Another important element is the R-loops that are detected at the *FMR1 locus* in PM cells and demethylated cells (Gerhardt, 2017)(Gerhardt, 2017)(Gerhardt, 2017)(Gerhardt, 2017). Although still unclear how the mechanism is, the presence of R-loops in the CpG Islands promoters seems to prevent DNA methylation and epigenetic silencing (Ginno et al., 2012)(Ginno et al., 2012)(Ginno et al., 2012)(Ginno et al., 2012). R-loops appear to favor the initiation of transcription increasing the likelihood that RNA polymerase binds to the promoter or facilitates the binding of additional transcription factors or chromatin modifiers to the promoter, which in turn promotes transcription (Usdin & Kumari, 2015)(Usdin & Kumari, 2015)(Usdin & Kumari, 2015)(Usdin & Kumari, 2015).

Besides that, in women with the PM allele, there is a higher presence of the histone marks H3K9ac (euchromatin-associated mark) and H3K9me2 (heterochromatin-associated mark) (Eslami et al., 2018)(Eslami et al., 2018)(Eslami et al., 2018)(Eslami et al., 2018). The H3K9ac mark is associated with active transcription (Gates et al., 2017)(Gates et al., 2017)(Gates et al., 2017)(Gates et al., 2017) and is strongly correlated with the over expression of the *FMR1* gene (Eslami et al., 2018)(Eslami et al., 2018)(Eslami et al., 2018)(Eslami et al., 2018).

#### 3.4.3. Epigenetic effects in Full Mutation (FM)

In most cases of FXS, the 5’ UTR end of *FMR1* is abnormally methylated and may inhibit the binding of transcription factors to the promoters of the gene (Tabolacci et al., 2005; Usdin & Kumari, 2015)(Tabolacci et al., 2005; Usdin & Kumari, 2015)(Tabolacci et al., 2005; Usdin & Kumari, 2015)(Tabolacci et al., 2005; Usdin & Kumari, 2015), since the chromatin associated with this region adopts a compacted conformation that is enriched with inactive chromatin markers (Kraan et al., 2018)(Kraan et al., 2018)(Kraan et al., 2018)(Kraan et al., 2018). In addition methylated CpG sites may be recognized by methyl radical binding proteins, such as MECP2 (Methyl-CpG Binding Protein 2) and MBD1-4 (Methyl-CpG Binding Domain Protein), which recruit deacetylases histone (HDACs) (Jones et al., 1998; Tabolacci et al., 2005)(Jones et al., 1998; Tabolacci et al., 2005)(Jones et al., 1998; Tabolacci et al., 2005)(Jones et al., 1998; Tabolacci et al., 2005). HDACs are enzymes responsible for removing the acetyl group from the lysines located in the tails of histones H3 and H4 (Seto & Yoshida, 2014; Tabolacci et al., 2005)(Seto & Yoshida, 2014; Tabolacci et al., 2005)(Seto & Yoshida, 2014; Tabolacci et al., 2005)(Seto & Yoshida, 2014; Tabolacci et al., 2005), making them less acetylated. Eventually, this leads to more condensed chromatin (Tabolacci et al., 2005; Usdin et al., 2014; Usdin & Kumari, 2015)(Tabolacci et al., 2005; Usdin et al., 2014; Usdin & Kumari, 2015)(Tabolacci et al., 2005; Usdin et al., 2014; Usdin & Kumari, 2015)(Tabolacci et al., 2005; Usdin et al., 2014; Usdin & Kumari, 2015).

Studies have shown that *FMR1* silencing in FM seems to occur in fetuses aged 8-12.5 weeks (Devys et al., 1992; Willemsen et al., 2002)(Devys et al., 1992; Willemsen et al., 2002)(Devys et al., 1992; Willemsen et al., 2002)(Devys et al., 1992; Willemsen et al., 2002). Although the sequence of events involved in the methylation of FM alleles is still unclear, FMRP appears to play an important role early in human neurological development (Sheridan et al., 2011)(Sheridan et al., 2011)(Sheridan et al., 2011)(Sheridan et al., 2011).

In FXS, FM alleles are associated with some histone marks, such as: H3-dimethylated on lysine 9 (H3K9me2) and H3 trimethylated on lysine 27 (H3K27me3), which are modified histones from inactive genes and have a broad distribution in the allele (Kumari and Usdin, 2010). High levels of dimethylation of lysine 9 (H3K9me2) and trimethylation of lysine 27 (H3K27me3) are observed while low levels of methylation in lysine 4 (H3K4, involved in activation) are observed (Eslami et al., 2018; Tabolacci et al., 2005; Usdin et al., 2014)(Eslami et al., 2018; Tabolacci et al., 2005; Usdin et al., 2014)(Eslami et al., 2018; Tabolacci et al., 2005; Usdin et al., 2014)(Eslami et al., 2018; Tabolacci et al., 2005; Usdin et al., 2014). Studies with FXS hESCs show that the enrichment of H3K9me2 occurs after the differentiation of these cells into neurons (Gerhardt, 2017)(Gerhardt, 2017)(Gerhardt, 2017)(Gerhardt, 2017).

In rare cases of FM but without the fragile X phenotype, ie, where gene silencing does not occur, *FMR1* is enriched for H3K9me2, but does not show CpG methylation (Usdin & Kumari, 2015)(Usdin & Kumari, 2015)(Usdin & Kumari, 2015)(Usdin & Kumari, 2015).

These alleles are also associated with elevated levels of histone H3 trimethylated on lysine 9 (H3K9me3) and histone H4 trimethylated on lysine 20 (H4K20me3). These marks are associated with the repression of transcription (https://epigenie.com/key-epigenetic-players/histone-proteins-and-modifications/histone-h4k20/) (Kumari & Usdin, 2010)(Kumari & Usdin, 2010)(Kumari & Usdin, 2010)(Kumari & Usdin, 2010). The distribution of histone modifications suggests that the onset of *FMR1* gene silencing begins in the CGG repeat, and is modulated by the molecules H3K9me3 and H4K20me3, which are enriched in the CGG repeat region, while the other histone modifications are more evenly distributed along the 5’ end of the gene (Usdin et al., 2014)(Usdin et al., 2014)(Usdin et al., 2014)(Usdin et al., 2014).

In this direction, it was recently investigated the methylation status (5-methylcytosine (5mC)) and hydroxymethylation (5-hydroxymethylcytosine (5hmC)) of the *FMR1 locus* in eight patients with FXS (Brasa et al., 2016)(Brasa et al., 2016)(Brasa et al., 2016)(Brasa et al., 2016). The 5mC mark was found almost exclusively as symmetric methylation of CpG, while 5hmC is distributed over the promoter and in other regions of transcriptionally active genes (Brasa et al., 2016; Tahiliani et al., 2009)(Brasa et al., 2016; Tahiliani et al., 2009)(Brasa et al., 2016; Tahiliani et al., 2009)(Brasa et al., 2016; Tahiliani et al., 2009). In the study by Brasa and collaborators (2016), it was identified that the loss of 5hmC and H4K20me1 is associated with the epigenetic silencing of *FRM1*. In addition, the authors found three regions of *FMR1* that demonstrate epigenetic changes. The first is a region surrounding the TSS (promoter/CpG island and the upstream part of intron 1) of *FMR1*, where hypermethylation is accompanied by a decrease in the active histone marks H3K4me2/H4K20me1 and enriched with repressive marks, mainly H3K9me3 and H3K9me2. The region that surrounds the TSS of antisense *FMR1AS* RNA and the region rich in alternative splicing (between introns 13 and 16) also represent epigenetic changes: both have reduced levels of 5mC (Brasa et al., 2016)(Brasa et al., 2016)(Brasa et al., 2016)(Brasa et al., 2016).

The *FMR1* transcript also promotes gene silencing when the mRNA CGG repeat binds in a complementary way to the *FMR1* (DNA) gene promoter, forming an RNA-DNA duplex (Colak et al., 2014; Nageshwaran & Festenstein, 2015)(Colak et al., 2014; Nageshwaran & Festenstein, 2015)(Colak et al., 2014; Nageshwaran & Festenstein, 2015)(Colak et al., 2014; Nageshwaran & Festenstein, 2015). In hECSs FXS, the absence of this duplex in the promoter avoids silencing; on the other hand, when present, they appear to activate pathways that induce epigenetic changes and silencing of *FMR1*(Gerhardt, 2017)(Gerhardt, 2017)(Gerhardt, 2017)(Gerhardt, 2017)(Gerhardt, 2017)(Gerhardt, 2017). *FMR1* mRNA may also be acting as a long non-coding RNAs by recruiting histone modifiers proteins such as PRC2 (Polycomb Repressive Complex), for example (Usdin & Kumari, 2015)(Usdin & Kumari, 2015)(Usdin & Kumari, 2015)(Usdin & Kumari, 2015)(Usdin & Kumari, 2015)(Usdin & Kumari, 2015).

Some studies seek treatment for FXS based on the removal of DNA methylation modifications and/or histones that maintain the locus of *FMR1* inactive (Kraan et al., 2018)(Kraan et al., 2018)(Kraan et al., 2018)(Kraan et al., 2018)(Kraan et al., 2018)(Kraan et al., 2018). Fragile X-lymphoblastoid cells treated with the DNA 5-aza-2-deoxycytidine (5-azadC) methylation inhibitor presented the transcription of the mutated *FMR1* gene reactivated (Chiurazzi et al., 1998)(Chiurazzi et al., 1998)(Chiurazzi et al., 1998)(Chiurazzi et al., 1998)(Chiurazzi et al., 1998)(Chiurazzi et al., 1998). After this treatment a proportion of 70 to 90% of the cells became unmethylated, but the efficiency of transcription reactivation was only 15 to 20% (Pietrobono et al., 2002)(Pietrobono et al., 2002)(Pietrobono et al., 2002). The acetylation levels are much lower in FXS, however, the treatment with 5-azadC induced an increase in histone acetylation the promoter regions and in the exon 1 of *FMR1* (Reines et al., 1999; Tabolacci et al., 2005)(Reines et al., 1999; Tabolacci et al., 2005)(Reines et al., 1999; Tabolacci et al., 2005).

Similarly, when using FXS-iPS cells (FXS induced pluripotent stem cells) to model this condition, the demethylating agent 5-azacytidine (5-azaC) was able to reactivate the *FMR1* gene expression levels in about 15% to 45% of the cells, depending on the amount administered. (Bar-Nur et al., 2012)(Bar-Nur et al., 2012)(Bar-Nur et al., 2012).

## 4. Concluding Remarks

Here we performed a SLR to unveil the mutation types underlying the causal *FMR1* gene of FXS. We show that point mutations, deletions, and mosaicisms involving the *FMR1* gene and without association with the CGG trinucleotide expansions also lead to a clinical phenotype that ranges from developmental delay and intellectual disability to fragile X characteristics, that includes ovarian problems as observed in premutated womens. Although such types of genetic variants are not often reported, its detection is limited mainly due to genetic investigations to FXS restrict to the CGG expansion in the *FMR1* gene. Thus, studies involving complete gene sequence may allow the identification of new genetic variants in patients with typical symptoms and characteristics of FXS, allowing them to demonstrate that even without presenting the CGG expansions a person can be positively diagnosed as a FXS individual. The detection of new atypical variants of FXS will allow us to expand our knowledge of distinct genetic variations underlying this syndrome and use them as novel genetic markers to correctly diagnose people with typical symptoms for this syndrome. Finally, it is also important to mention that the non-CGG expansion was never considered definitely pathogenic, but only rare (in population dataset as controls) and without concluding evidence of clinical effect.

## Data Availability

All data produced in the present work are contained in the manuscript.

