## Supplementary data for "Non-CGG trinucleotide repeat expansions as pathogenic genetic mutations in Fragile X Syndrome"

**Suplementary Data**

**Table S1.** Search strategy used to perform SLR in the PubMed, Scopus and BVS databases.

| Data base | Search Parameters |
|---|---|
| PubMed | (Xq27 [Title/Abstract] OR Fragile X Mental Retardation 1[Title/Abstract] OR Fragile X Mental Retardation Protein 1[Title/Abstract] OR FMRP[Title/Abstract] OR Synaptic Functional Regulator FMR1[Title/Abstract] OR Premature Ovarian Failure 1[Title/Abstract] OR Protein FMR-1[Title/Abstract] OR FRAXA[Title/Abstract] OR POF1[Title/Abstract] OR POF[Title/Abstract] OR FXTAS[Title/Abstract]) AND (variant[Title/Abstract] OR Polimorphism[Title/Abstract] OR alteration[Title/Abstract] OR mutation[Title/Abstract] OR change[Title/Abstract] OR SNP[Title/Abstract] OR deletion[Title/Abstract] OR insertion[Title/Abstract] OR nonsense[Title/Abstract] OR missense[Title/Abstract] OR SNV[Title/Abstract] OR duplication[Title/Abstract] OR Frameshift[Title/Abstract] OR silent[Title/Abstract] OR expression[Title/Abstract] OR modulation[Title/Abstract]) AND (gene[Title/Abstract] OR Transcript[Title/Abstract] OR Transcriptomic[Title/Abstract] OR mRNA[Title/Abstract] OR RNA[Title/Abstract] OR Messenger RNA[Title/Abstract] OR DNA[Title/Abstract] OR Genome[Title/Abstract] OR Genetic[Title/Abstract] OR Genomic[Title/Abstract] OR Protein[Title/Abstract] OR proteomic[Title/Abstract] OR aminoacid[Title/Abstract]) |
| Scopus | TITLE-ABS-KEY ( xq27 ) OR TITLE-ABS-KEY ( fragile AND x AND mental AND retardation 1 ) OR TITLE-ABS-KEY ( fragile AND x AND mental AND retardation AND protein 1 ) OR TITLE-ABS-KEY ( fmrp ) OR TITLE-ABS-KEY ( synaptic AND functional AND regulator AND fmr1 ) OR TITLE-ABS-KEY ( premature AND ovarian AND failure 1 ) OR TITLE-ABS-KEY ( fxtas ) OR TITLE-ABS-KEY ( protein AND fmr-1 ) OR TITLE-ABS-KEY ( pof1 ) OR TITLE-ABS-KEY ( pof ) AND TITLE-ABS-KEY ( variant ) OR TITLE-ABS-KEY ( polimorphism ) OR TITLE-ABS-KEY ( alteration ) OR TITLE-ABS-KEY ( mutation ) OR TITLE-ABS-KEY ( change ) OR TITLE-ABS-KEY ( snp ) OR TITLE-ABS-KEY ( snv ) OR TITLE-ABS-KEY ( deletion ) OR TITLE-ABS-KEY ( insertion ) OR TITLE-ABS-KEY ( nonsense ) OR TITLE-ABS-KEY ( missense ) OR TITLE-ABS-KEY ( duplication ) OR TITLE-ABS-KEY ( frameshift ) OR TITLE-ABS-KEY ( silent ) OR TITLE-ABS-KEY ( expression ) OR TITLE-ABS-KEY ( modulation ) AND TITLE-ABS-KEY ( gene ) OR TITLE-ABS-KEY ( transcript ) OR TITLE-ABS-KEY ( transcriptomic ) OR TITLE-ABS-KEY ( mrna ) OR TITLE-ABS-KEY ( rna ) OR TITLE-ABS-KEY ( messenger AND rna ) OR TITLE-ABS-KEY ( dna ) OR TITLE-ABS-KEY ( genome ) OR TITLE-ABS-KEY ( genetic ) OR TITLE-ABS-KEY ( genomic ) OR TITLE-ABS-KEY ( protein ) OR TITLE-ABS-KEY ( proteomic ) OR TITLE-ABS-KEY ( aminoacid) |
| BVS | ("Xq27" OR "Fragile X Mental Retardation 1" OR "Fragile X Mental Retardation Protein 1" OR "FMRP" OR "Synaptic Functional Regulator FMR1" OR "Premature Ovarian Failure 1" OR "Protein FMR-1" OR "FRAXA" OR "POF1" OR "POF" OR "FXTAS") AND ("variant" OR "Polimorphism" OR "alteration" OR "mutation" OR "change" OR "SNP" OR "deletion" OR "insertion" OR "nonsense" OR "missense" OR "SNV" OR "duplication" OR "Frameshift" OR "silent" OR "expression" OR "modulation") AND ("gene" OR "Transcript" OR |

| | |
|---|---|
| | "Transcriptomic" OR "mRNA" OR "RNA" OR "Messenger RNA" OR "DNA" OR "Genome" OR "Genetic" OR "Genomic" OR "Protein" OR "proteomic" OR "aminoacid") |

**Table S2. Revision of the previously identified variants in the *FMR1* gene of individuals with developmental delay and/or intellectual disability.** Reported variants follow the coordinates of the human reference genome, build GRCH38. Variant: correspond to the genetic variant found; Molecular consequence: refers to the type of mutation and its effect on the chemical constitution of the gene; Protein effect: alteration in the amino acids of the protein; Gene region: describes the *FMR1* region where the variant was described; *Locus*: genomic location of the genetic variant found, based on the GRCh38 reference genome; Isoform: refers to different mRNAs that encode multiple FMRP proteins; Phenotype: characteristics / symptoms observed in the individual with the genetic variant; Clinical significance: pathogenicity of variants; Reference: individual list of all articles consulted.

| **Variant** | **Molecular consequence** | **Protein Effect** | **Gene region** | ***Locus*** | **Isoform** | **Phenotype** | **Clinical significance** | **Reference** |
|---|---|---|---|---|---|---|---|---|
| c.-413C>G | Non-coding | - | Promoter | chrX:147,911,767 | NM_002024 | Intellectual disability | Uncertain significance | (Grasso et al., 2010; Milà et al., 2000) |
| c.-332G>C | Non-coding | - | Promoter | chrX:147,911,848 | NM_002024 | Developmental delay | Uncertain significance | (Collins et al., 2010a) |
| c.-293T>C | Non-coding | - | Promoter | chrX:147,911,887 | NM_002024 | Developmental delay | Uncertain significance | (Collins et al., 2010b) |
| c.-254A>G | Non-coding | - | Promoter | chrX:147,911,926 | NM_002024 | Developmental delay | Uncertain significance | (Collins et al., 2010b) |

| | | | | | | | | |
|---|---|---|---|---|---|---|---|---|
| c.18G>T | Synonymous | p.Val6= | exon 1 | chrX:147,912,197 | NM_002024 | Developmental delay | Benign | (Collins et al., 2010b) |
| c.105-8A>C | Non-coding | - | intron 2 | chrX:147,925,532 | NM_002024 | Developmental delay | Uncertain significance | (Collins et al., 2010b) |
| c.377T>C [1] | Nonsynonymous | p.Phe126Ser | exon 5 | chrX:147,928,765 | NM_002024 | Developmental disorders | Likely pathogenic | (Wright et al., 2015a) |
| c.413G>A | Nonsynonymous | p.Arg138Gln | exon 5 | chrX:147,928,801 | NM_002024 | Intellectual disability, developmental delay and seizures | Uncertain significance | (Collins et al., 2010b; Myrick et al., 2015) |
| c.630+438A>C | Non-coding | - | intron 7 | chrX:147,930,682 | NM_002024 | Developmental delay | Uncertain significance | (Collins et al., 2010b) |
| c.631-840G>A | Non-coding | - | intron 7 | chrX:147,931,585 | NM_002024 | Developmental delay | Uncertain significance | (Collins et al., 2010b) |
| c.677G>A | Nonsynonymous | p.Arg226Lis | exon 8 | chrX:147,932,471 | NM_001185082; NM_001185076 | Moderate to severe intellectual disability | Likely pathogenic | (Grozeva et al., 2015a) |
| c.767A>G | Nonsynonymous | p.Asp256Gli | exon 8 | chrX:147,932,561 | NM_001185082; NM_001185076 | Moderate to severe intellectual disability | Likely Benign | (Grozeva et al., 2015b) |
| c.880+885A>G | Non-coding | - | intron 9 | chrX:147,933,648 | NM_002024 | Developmental delay | | (Collins et al., 2010b) |
| c.990+4T>C | Non-coding | - | intron 10 | chrX:147,936,617 | NM_002024 | Developmental delay | Uncertain significance | (Collins et al., 2010b) |
| c.1189-39A>G | Non-coding | - | intron 12 | chrX:147,940,537 | NM_002024 | Intellectual disability and | non-pathogenic | (Handt et al., 2014a) |

[1] Genetic findings reported by Quartier *et al.*, 2017.

| | | | | | | | | |
|---|---|---|---|---|---|---|---|---|
| | | | | | | developmental delay | | |
| c.1168G>A | Nonsynonymous | p.Ala390Tre | exon 12 | chrX:147,940,618 | NM_001185082; NM_001185076 | Moderate to severe intellectual disability | Likely Benign | (Grozeva et al., 2015b) |
| c.1325G>A [2] | Nonsynonymous | p.Arg442Gln | exon 14 | chrX:147,943,180 | NM_002024 | Developmental disorders | Uncertain significance | (Mangano et al., 2022; Wright et al., 2015b; Zeidler et al., 2021) |
| c.1444G>A | Nonsynonymous | p.Gly482Ser | exon 14 | chrX:147,943,299 | NM_002024 | Intellectual disability and developmental delay | Uncertain significance | (Handt et al., 2014b) |
| c.1472-521C>G | Non-coding | - | intron 14 | chrX:147,944,348 | NM_002024 | Developmental delay | Uncertain significance | (Collins et al., 2010b) |
| c.1601G>A | Nonsynonymous | p.Arg534His | exon 15 | chrX:147,944,998 | NM_002024 | Intellectual disability and developmental delay | Uncertain significance | (Handt et al., 2014b) |
| c.1618G>A | Nonsynonymous | p.Gly540Glu | exon 15 | chrX:147,945,015 | NM_002024 | Intellectual disability | Uncertain significance | (Hu et al., 2016) |
| c.1301C>T | Nonsynonymous | p.Ala434Val | exon 14 | chrX:147,945,032 | NM_001185075; NM_001185081 | Moderate to severe intellectual disability | Likely Benign | (Grozeva et al., 2015b) |
| c.1586C>T | Nonsynonymous | p.Tre529Ile | exon 16 | chrX:147,948,802 | NM_001185075; NM_001185081 | Moderate to severe intellectual disability | Likely pathogenic | (Grozeva et al., 2015b) |

[2] Genetic findings reported by Quartier *et al.*, 2017.

| | | | | | | | | |
|---|---|---|---|---|---|---|---|---|
| c.1610C>T | Nonsynonymous | p.Ser537Leu | exon 16 | chrX:147,948,826 | NM_001185075; NM_001185081 | Moderate to severe intellectual disability | Likely pathogenic | (Grozeva et al., 2015b) |
| c.*23T>C | Non-coding | - | 3'UTR | chrX:147,948,867 | NM_002024 | Developmental delay | Uncertain significance | (Collins et al., 2010b) |
| c.*60G >*C* | Non-coding | - | 3'UTR | chrX:147,948,904 | NM_002024 | Intellectual disability and developmental delay | non-pathogenic | (Handt et al., 2014b) |
| c.*746T>C | Non-coding | - | 3'UTR | chrX:147,948,912 | NM_002024 | Developmental delay | Uncertain significance | (Collins et al., 2010b; Suhl et al., 2015) |
| c.*68T >*C* | Non-coding | - | 3'UTR | chrX:147,948,912 | NM_002024 | Intellectual disability and developmental delay | non-pathogenic | (Handt et al., 2014b) |
| c.*1867G>A | Non-coding | - | 3'UTR | chrX:147,950,711 | NM_002024 | Developmental delay | Uncertain significance | (Collins et al., 2010b) |
| c.*2035C>T | Non-coding | - | 3'UTR | chrX:147,950,879 | NM_002024 | Developmental delay | Uncertain significance | (Collins et al., 2010b) |
| c.1256C>T | Nonsynonymous | - | exon 14 | chrX:147,944,987 | NM_001185081 | Intellectual disability and developmental delay | Uncertain significance | (Xu et al., 2021) |
| c.881-1G>T | Nonsynonymous | - | exon 10 | chrX:147,936,503 | NM_002024 | Intellectual disability | Likely pathogenic | (Carroll et al., 2020) |
| c.801G>A | Synonymous | p.Glu267Glu | exon 8 | chrX:147,932,595 | NM_002024 | Intellectual disability and developmental delay | Likely pathogenic | (Carion et al., 2020) |
| c.1216C>T | Stopgain | p.Gln541Ser | exon 15 | chrX:147,945,018 | NM_002024 | Intellectual disability | Likely pathogenic | (Park et al., 2020) |

**Table S3. Review of the previously identified variants of the *FMR1* gene of individuals with the typical FXS phenotype.** Reported variants follow the coordinates of the human reference genome, build GRCH38. Variant: correspond to the genetic variant found; Molecular consequence: refers to the type of mutation and its effect on the chemical constitution of the gene; Protein effect: alteration in the amino acids of the protein; Gene region: describes the *FMR1* region where the variant was described; *Locus*: genomic location of the genetic variant found, based on the GRCh38 reference genome; Isoform: refers to different mRNAs that encode multiple FMRP proteins; Phenotype: characteristics/symptoms observed in the individual with the genetic variant; Clinical signifance: pathogenicity of variants; Reference: individual list of all articles consulted.

| **Variant** | **Molecular consequence** | **Protein Effect** | **Gene region** | ***Locus*** | **Isoform** | **Phenotype** | **Clinical significance** | **Reference** |
|---|---|---|---|---|---|---|---|---|
| c.52-1_52delinsTA | Stopgain | p.(?) | exon 2 | chrX:147,921,932-147,921,933 | NM_002024 | Dysmorphic features, elongated face, forehead and prominent ears, macroorchidia, hyperextensibility of the joints, autistic characteristics and developmental delay. | Pathogenic | (Lugenbeel et al., 1995a) |
| c.80C>A | Stopgain | p.Ser27* | exon 2 | chrX:147,921,961 | NM_002024 | Intellectual deficiency, facial dysmorphism, macroorchidism, epilepsy, autistic characteristics and little use of language. | Pathogenic | (Gronskov et al., 2011) |

| | | | | | | | | |
|---|---|---|---|---|---|---|---|---|
| c.373delA [3] | Frameshift | p.Thr125Leufs*35 | exon 5 | chrX:147,928,761 | NM_002024 | Dysmorphic features, elongated face, forehead and prominent ears, hyperextensibility of joints, autistic characteristics, ADHD, developmental delay and language impairment. | Pathogenic | (Lugenbeel et al., 1995b) |
| c.413G>A | Nonsynonymous | p.Arg138Gln | exon 5 | chrX:147,928,801 | NM_002024 | Developmental delay, autistic behavior, attention deficit hyperactivity disorder, seizures, handshake, elongated face, large ears, soft hands, hyperextensible fingers and flat feet | Likely pathogenic | (Sitzmann et al., 2018) |
| c.420-8A>G | Frameshift | p.Met140Ilefs*3 | intron 5 | chrX:147,929,940 | NM_002024 | Delay in language, elongated face, stereotypies and repetitive behaviors, with hand shaking and anxiety. | Pathogenic | (Quartier et al., 2017a) |
| c.797G>A | Nonsynonymous | p.Gly266Glu | exon 8 | chrX:147,932,591 | NM_002024 | Delayed development, along with multiple other behaviors and physical characteristics commonly associated with SXF | Likely pathogenic | (Myrick et al., 2014) |
| c.911T>A [4] | Nonsynonymous | p.Ile304Asn | exon 10 | chrX:147,936,534 | NM_002024 | Developmental delay, elongated face, large ears, prominent jaw and macroorchidism. | Pathogenic | (De Boulle et al., 1993) |
| c.990+1G>A | Frameshift | p.Lys295Asnfs * 11 | intron 10 | chrX:147,936,614 | NM_002024 | Delayed motor skills and delayed verbal communication. Features repetitive behavior, as well as shake hands and avoid physical contact. Tall forehead, coarse face, wide mouth, | Pathogenic | (Quartier et al., 2017b) |

[3] Genetic findings reported by Quartier *et al.*, 2017.

[4] Genetic findings reported by Quartier *et al.*, 2017.

| | | | | | | | | |
|---|---|---|---|---|---|---|---|---|
| | | | | | | full-sized ears but abnormal shape. Autistic traits. | | |
| c.1637 G>A | Nonsynonymous | p.Arg546His | exon 15 | chrX:147,945,034 | NM_002024 | Intellectual disability, elongated face, prominent jaw and large forehead. | Uncertain significance | (Quartier et al., 2017b; Wang et al., 1997) |
| c.1457 insG | Frameshift | p.Gly538fs*23 | exon 15 | chrX:147,945,607 | NM_001185075 | Physical and behavioral characteristics typical of FXS and moderate to severe intellectual disability. | Uncertain significance | (Okray et al., 2015) |
| c.25_131=/del | Frameshift | - | exon 1 | chrX:147,911,943-147,912,049 | NM_001185081.2 | Aggressive behaviour, limited language skills, sleep disturbance, hyperactivity and problems with attention (ADHD), and severe anxiety, broad and wide palate, slight clinodactyly of the fifth finger of his right hand, and hyper-extensible finger joints. | Uncertain significance | (Winarni *et al.*, 2025) |
| c.25_131=/del | Frameshift | - | exon 1 | chrX:147,911,943-147,912,049 | NM_001185081.2 | Self-harming and aggressive behaviour, developmental delay and delayed communication skills, sleep disturbance and hyperactivity and attention (ADHD), and separation anxiety, repetitive speech and other stereotypic behavior. | Uncertain significance | (Winarni *et al.*, 2025) |

| | | | | | | | | |
|---|---|---|---|---|---|---|---|---|
| c.-130_183del | Non-coding | - | promoter | chrX:147,911,737-147,912,049 | NM_001185081.2 | Mild FXS features, school performance below average . | Likely pathogenic | (Hnoonual *et al*., 2024) |
| c.1_4107del | Full deletion of gene | - | exon 1 - 17 | chrX:146,990,647-147,058,715 | NM_002024 | ID, emotional lability, certain shyness with little habitual contact, open mouth with frequent drooling, elongated face with a large forehead and large and low-set pinnae, joint hypermobility in the upper limbs and subtle kyphosis. | Pathogenic | (Gómez-Rodrigues *et al*., 2022) |
