## Supplementary material for "Non-CGG trinucleotide repeat expansions as pathogenic genetic mutations in Fragile X Syndrome": Tables and table legends

### Tables and tables legends

**Table 1 – Molecular consequence of the previously reported variants in the *FMR1* gene.** Variant: correspond to the genetic variant found; Molecular consequence: refers to the type of mutation and its effect on the chemical constitution of the gene; Protein effect: alteration in the amino acids of the protein.

| Variant | Molecular consequence | Protein effect |
|---|---|---|
| c.-413C>G | Non-coding | - |
| c.-332G>C | Non-coding | - |
| c.-293T>C | Non-coding | - |
| c.-254A>G | Non-coding | - |
| c.18G>T | Synonymous | p.Val6= |
| c.105-8A>C | Non-coding | - |
| c.377T>C | Nonsynonymous | p.Phe126Ser |
| c.413G>A | Nonsynonymous | p.Arg138Gln |
| c.630+438A>C | Non-coding | - |
| c.631-840G>A | Non-coding | - |
| c.677G>A | Nonsynonymous | p.Arg226Lis |
| c.767A>G | Nonsynonymous | p.Asp256Gli |
| c.880+885A>G | Non-coding | - |
| c.990+4T>C | Non-coding | - |
| c.1189-39A>G | Non-coding | - |
| c.1168G>A | Nonsynonymous | p.Ala390Tre |
| c.1325G>A | Nonsynonymous | p.Arg442Gln |
| c.1444G>A | Nonsynonymous | p.Gly482Ser |
| c.1472-521C>G | Non-coding | - |
| c.1601G>A | Nonsynonymous | p.Arg534His |
| c.1618G>A | Nonsynonymous | p.Gly540Glu |
| c.1301C>T | Nonsynonymous | p.Ala434Val |
| c.1586C>T | Nonsynonymous | p.Tre529Ile |
| c.1610C>T | Nonsynonymous | p.Ser537Leu |
| c.*23T>C | Non-coding | - |
| c.*60G > *C* | Non-coding | - |
| c.*746T>C | Non-coding | - |
| c.*68T > *C* | Non-coding | - |
| c.*1867G>A | Non-coding | - |
| c.*2035C>T | Non-coding | - |
| c.52-1_52delinsTA | Stopgain | p.(?) |
| c.80C>A | Stopgain | p.Ser27* |
| c.373delA | Frameshift | p.Thr125Leufs*35 |
| c.420-8A>G | Frameshift | p.Met140Ilefs*3 |
| c.797G>A | Nonsynonymous | p.Gly266Glu |
| c.911T>A | Nonsynonymous | p.Ile304Asn |
| c.990+1G>A | Frameshift | p.Lys295Asnfs*11 |
| c.1637G>A | Nonsynonymous | p.Arg546His |
| c.1457insG | Frameshift | p.Gly538fs*23 |
| c.1256C>T | Nonsynonymous | - |
| c.881-1G > T | Nonsynonymous | - |
| c.801G > A | Nonsynonymous | p.Glu267Glu |
| c.1216C>T | Stopgain | p.Gln406Ser |

**Table 2 – Relation of the cases of mosaicisms found in the literature involving deletions and associated with SXF**. Mosaic, refers to the type of mosaic; Genotype, refers to the genetic makeup of mosaicism; Phenotype, refers to the condition/symptoms observed in the individual with mosaicism; Reference, refers to the individual list of all the works consulted.

| Mosaic | Genotype | Phenotype | Reference |
|---|---|---|---|
| Mo:MCDel | Presence of CM (6.5 kb) and deletion of ~ 100 bp | Clinical phenotype of FXS. The deletion did not impair the transcription and translation of FMRP, which was present in 28% of the individual lymphocytes. | (de Graaff et al., 1996) |
| Mo:MCDel | Presence of methylated CM (300 to 350 CGG repeats) and 905 bp deletion. This deletion encompasses the CGG repeats and excludes a start codon (ATG). | FXS with absence of FMRP protein, since neither the CM allele nor the deletion could be transcribed. | (Arocena et al., 2000) |
| Mo:MCMePMDel | Presence of CM (6-8 kb, 10%), PM (3.2 kb) and unmethylated CM (3.5 kb, 37%) and a deletion (less than 2.8 kb, 53%) covering the entire region of the CGG repeats 42bb upstream of it. | Atypical X Fragile, with learning disabilities, but without mental retardation. FMRP is expressed in only 22% of cells and the level of mRNA is increased by 3.6 fold. | (Han et al., 2006) |
| Mo:MCPMDel | Presence of 2 methylated CM alleles (230 and 450 CGGs), one PM allele (170 CGGs, 25%) and a 175bp deletion (15%). | Individual of a classical SXF family, but without the characteristic phenotype of SXF. Expression of FMRP in lymphocytes was 18%. | (Govaerts et al., 2007) |
| Mo:Ndel | Presence of normal allele (23 CGGs) and deletion of 1,013,395bp, being responsible for the deletion of all *FMR1* gene and its neighbor *FMR1NB* gene. The deletion was present in 90% of the patient's lymphocytes and the normal allele in 10%. | Cognitive and behavioral profile with characteristics compatible with SXF. | (Coffee et al., 2008) |
| Mo:MCMe | mosaic of unmethylated MC size (CGG repeat size ranging from 180 to 410) | FXTAS with *FMR1* mRNA level 7 times higher than the normal mean and reduced FMRP expression levels (38% of normal). | (Santa María et al., 2014) |
| Germinal mosaic | Deletion of 300 kb in Xq27.3 spanning the entire *FMR1* gene | Mosaicism found in the asymptomatic mother of a boy with FXS: delayed physical and linguistic development and autism spectrum disorder. | (Jiraanont et al., 2016) |
| Mo:MCMeDel | Presence of unmethylated and methylated CM alleles and a microdeletion involving a ~ 80 bp sequence in the FMR1 promoter, as well as complete loss of the CGG repeat in part of the cells. | FXS and clinical features compatible with FXTAS. | (Hwang et al., 2016) |
| Mo:MCMeDel | Presence of MC alleles (methylated and unmethylated) and with one allele covering a CGG repeat deletion present in 13% of the cells. The total percentage of cells carrying hypermethylated full mutation alleles was approximately 60%. | ADHD, anxiety, finger bite, poor eye contact, palpitation of hands, long face and prominent ears | (Jiraanont et al., 2017) |

Abreviations: Mo:MCDel, mosaicism for complete mutation and deletion; Mo:MCMePMDel, mosaicism for complete mutation, methylation state, pre-mutation and deletion; Mo:MCPMDel, mosaicism for complete mutation, pre-mutation, and deletion; Mo:Ndel, mosaicism for normal allele and deletion; Mo:MCMe, mosaicism for complete mutation and methylation state; Mo:MCMeDel, mosaicism for complete mutation, state of methylation and deletion; FXS, Fragile X Syndrome; CM, complete mutation; PM, Pre-mutation; FXTAS, Fragile X-associated tremor/ataxia syndrome.
